# Demographic profiles of contact lens wearers and their association with lens wear characteristics in Indonesia: A cross-sectional study

**DOI:** 10.64898/2026.09.09.26362601

**Authors:** Tri Rahayu, Nabila Aljufri, Umar Mardianto, Angelina Patricia Chandra, Agus Sugiharto

## Abstract

**Introduction:** Contact lens (CL) use is increasing globally, yet epidemiological data on CL wearers remain limited in Southeast Asia, including Indonesia. This study aimed to describe the demographic characteristics of CL users, the distribution of CL types, and the factors associated with CL wear patterns and correction status in Indonesia.

**Methods:** This cross-sectional study used convenience sampling to recruit current CL users aged 18–40 years through online advertisements and an ophthalmology clinic in Indonesia. Participants self-reported demographic data, CL type, reasons for use, and refractive correction details. Associations between categorical variables were assessed using chi-square or Fisher’s exact tests. Agreement between recommended and actual CL spherical power was evaluated using the Wilcoxon signed-rank test, and factors associated with correction status were analyzed using multinomial logistic regression.

**Results:** Among 350 participants, the majority were female (95.4%), aged 18–30 years (54.0%), held a bachelor’s degree (66.6%), and earned ≥Rp5,000,000 monthly (64.9%). Soft colored lenses were the most commonly used type (60.1%), and practicality was the most frequently cited reason for use (38.1%), followed by cosmetic purposes (31.1%) and refractive correction (30.8%). Mild myopia was the most prevalent refractive error (40.0%). Undercorrection was the most common CL correction status (44.4% OD, 44.8% OS), with actual CL power significantly less negative than the recommended power in both eyes (OD: −0.18 D, p=0.015; OS: −0.24 D, p=0.001). Greater myopia severity was significantly associated with both undercorrection (OR 3.63–5.01) and overcorrection (OR 4.22–5.83) in both eyes (all p<0.001), while age, wear duration, and sex showed no significant association.

**Conclusion:** This first nationwide demographic study of CL wearers in Indonesia found a predominance of colored soft lenses and a substantial tendency toward undercorrection linked to myopia severity. These findings highlight the need for public education on professional CL fitting, prescribing, and safe use practices.

## Introduction

Contact lenses (CLs) are widely used medical devices designed to rest on the anterior surface of the eye to correct refractive errors by replacing or complementing the optical properties of the cornea.^[1]^ Beyond refractive correction, contact lenses have important therapeutic, preventive, diagnostic, cosmetic, and occupational applications, making them an integral component of modern eye care.^[2]^ Advances in lens materials and designs have improved oxygen permeability, comfort, optical performance, and safety, leading to the development of various lens types, including soft contact lenses, rigid gas-permeable (RGP) lenses, and specialized designs such as orthokeratology (Ortho-K), multifocal, and cosmetic contact lenses.^[3]^

Globally, contact lens use continues to increase, with an estimated 140 million wearers worldwide.^[4]^ Technological innovations over the past decade have expanded the range of available lens materials and replacement modalities, influencing both prescribing practices and wearer preferences.^[5]^ Soft contact lenses prescribed for daily wear remain the most commonly used modality globally, with women comprising the majority of contact lens wearers.^[6–10]^ Moreover, the adoption of silicone hydrogel daily wear lenses worldwide has also increased substantially in recent years.^[11]^ In contrast, rigid contact lenses account for only a small proportion of lens fittings, particularly in developing countries.^[12]^ Contact lens prescribing and wear are influenced by multiple factors, including demographic characteristics, socioeconomic status, indications for lens wear, the prevalence of ocular conditions, availability of contact lens options, practitioner expertise, and sociocultural influences.^[8,13]^ The prevalence of contact lens wear and prescribing patterns differ considerably between countries due to variations in demographic characteristics, socioeconomic conditions, cultural preferences, access to eye care services, and the availability of different contact lens modalities.^[14]^ More recent evidence from Japan and Taiwan similarly demonstrates the predominance of soft contact lenses, including increasing adoption of silicone hydrogel and daily disposable lenses, reflecting ongoing technological advancements and changing wearer preferences.^[14]^

Despite the rapid growth of the contact lens market in Asia,^[15]^ epidemiological data remain limited in several Southeast Asian countries, including Indonesia. Given the country’s large population, increasing prevalence of refractive errors,^[16,17]^ and expanding access to eye care services, understanding local contact lens use is essential. In Indonesia, contact lenses are widely available through optical stores, beauty retailers, and online marketplaces, and their purchase is generally not restricted to prescriptions issued by ophthalmologists or other eye care professionals. Consequently, many individuals are able to obtain contact lenses without a formal eye examination or professional fitting. Therefore, this study aimed to describe the characteristics of contact lens users, the distribution of contact lens types in Indonesia, and the demographic factors associated with different patterns of lens wear.

## Methods

### Ethical considerations

Ethical approval was obtained from the Health Research Ethics Committee, Faculty of Medicine, Universitas Indonesia (Reference No. Ket-1196/UN2.F1/ETIK/PPM.00.01/2024).

### Study design

This cross-sectional study recruited volunteer participants in Indonesia using a convenience sampling approach for the purpose of validating the quality of contact lens wearer questionnaire in another study. Individuals who were current or previous contact lens users were invited to participate through online advertisements on social media platforms and in-person recruitment through an ophthalmology clinic. Eligible participants who voluntarily agreed to participate were provided with detailed information regarding the study objectives and procedures before giving written or electronic informed consent.

### Inclusion and exclusion criteria

Eligible participants were individuals aged 18–40 years who were current contact lens users and voluntarily agreed to participate in the study. Participants who submitted the questionnaire after the predefined data collection period had ended were excluded from the analysis. In addition, responses that were incomplete, duplicated, or contained insufficient information for analysis were excluded where applicable.

### Data collection procedure

Participants were asked to self-report the refractive correction prescribed for each eye, including the spherical power (D) and cylindrical power (D) from their most recent spectacle or contact lens prescription. The spherical equivalent (SE) was calculated as the spherical power plus one-half of the cylindrical power. Myopia was defined as a SE of ≤ −0.50 diopters (D) for each eye. Based on SE, myopia was categorized as mild (−0.50 to −3.00 D), moderate (−3.01 to −5.99 D), and high (< −6.00 D).^[18]^ Astigmatism severity was classified according to cylindrical power as mild (0.50 to <1.50 D), moderate (1.50 to <3.00 D), and high (≥3.00 D).^[19]^

To ensure data quality, responses were excluded if refractive values could not be interpreted reliably. These included prescriptions in which the sign (+/−) of the spherical power was omitted, responses indicating no refractive error or normal vision, entries containing only a dash (“-”), or prescriptions with inconsistent notation between the right and left eyes (e.g., one eye recorded as a negative value while the other lacked a sign, making the refractive status ambiguous). When only a single refractive value was provided, it was assumed to represent the spherical power with a negative sign, as participants commonly omitted the minus symbol when reporting myopic prescriptions. Participants with incomplete or uninterpretable refractive data were excluded from analyses involving spherical equivalent.

### Statistical analysis

Statistical analyses were performed using IBM SPSS Statistics version 27.0 (IBM Corp., Armonk, NY, USA). Continuous variables were summarized as mean ± standard deviation (SD), while categorical variables were presented as frequencies and percentages.

For the analysis of contact lens type and reasons for contact lens use according to sex and age group, Pearson’s chi-square test was used to assess associations between categorical variables. Fisher’s exact test was used when the expected cell count was less than five. For analyses involving multiple-response variables, each response category was analyzed independently and percentages were calculated based on the total number of participants; therefore, percentages could exceed 100%.

Agreement analysis was performed among all participants with refractive error. For participants with spherical equivalent refraction of ≥ ±4.00 diopters (D), the spherical power was converted to account for the difference in vertex distance between spectacle and contact lens correction. Spectacle refraction is measured at a vertex distance of approximately 12 mm, whereas contact lenses are positioned directly on the corneal surface (vertex distance = 0 mm). Therefore, vertex distance compensation was applied to spherical powers of ≥ ±4.00 D, for which the effect of vertex distance is considered clinically relevant. For spherical powers below ±4.00 D, no vertex distance conversion was applied because the resulting change in effective power is generally <0.25 D and is considered clinically insignificant. Accordingly, negative spherical powers of ≥ −4.00 D were adjusted by adding 0.25 D (e.g., −4.00 D was converted to −3.75 D), whereas positive spherical powers of ≥ +4.00 D were adjusted by adding 0.25 D (e.g., +4.00 D was converted to +4.25 D).^[20]^

The recommended contact lens spherical power was calculated by converting the spectacle spherical equivalent refraction power using vertex distance compensation. Correction status was subsequently classified as appropriate correction, undercorrection, or overcorrection by comparing the recommended spherical power with the participant-reported contact lens spherical power. Differences between the recommended and actual contact lens spherical powers were analyzed separately for the right (OD) and left (OS) eyes using the Wilcoxon signed-rank test, as the paired differences were not normally distributed. A two-sided *p* value <0.05 was considered statistically significant.

Factors associated with contact lens correction status were evaluated using multinomial logistic regression, performed separately for the right and left eyes. Correction status was entered as the dependent variable, with appropriate correction as the reference category. The independent variables included age group, duration of contact lens wear, myopia severity, and sex. The reference categories were age 18–30 years, contact lens wear duration <1 year, and female sex, respectively. Results were presented as odds ratios (ORs) with 95% CIs and p-values. An OR >1 indicated higher odds of belonging to the specified correction-status category relative to appropriate correction, whereas an OR <1 indicated lower odds. Model significance was assessed using the likelihood-ratio chi-square test, and model fit was summarized using Nagelkerke’s R². Statistical significance was defined as a two-sided p-value <0.05.

## Results

### Participant characteristics

A total of 350 participants were included in this study. More than half were aged 18–30 years (54.0%) with the mean age of 29.52 ± 4.31, and the sample was predominantly female (95.4%), with only 4.6% male participants. Most participants held a bachelor’s degree (66.6%), followed by a postgraduate degree (20.6%), high school or equivalent education (7.7%), and a diploma (5.1%). In terms of employment, private-sector employees comprised the largest group (35.4%), followed by doctors (19.1%), students (13.1%), housewives (11.4%), and entrepreneurs (11.4%), with smaller proportions working as healthcare professionals, teachers, or in other occupations. Most participants were married (57.7%), while 40.9% were single and 1.4% were divorced. Regarding monthly income, 64.9% reported earning more than Rp5,000,000, 21.4% earned less than Rp3,000,000, and 13.7% earned between Rp3,000,000 and Rp5,000,000. The predominance of participants with a monthly income of ≥Rp5 million (64.9%) suggests that contact lens use in this study was more common among individuals with relatively higher socioeconomic status. This finding may reflect the financial requirements associated with contact lens wear, including the initial cost of lenses as well as recurring expenses for replacement, lens care products, and related eye-care services. In 2026, provincial minimum wages in Indonesia range from approximately Rp2.31 million to Rp5.72 million per month, with substantial variation between regions and additional variation at the city level.^[21]^ Thus, an income of ≥Rp5 million generally represents an income at or above the upper range of provincial minimum wages in Indonesia. This socioeconomic pattern may also be related to the geographic distribution of participants in this study, of whom 76.9% were from Java. Java contains several of Indonesia’s major urban and economic centers, where income levels, access to eye-care services, and availability of optical products may be relatively higher. Therefore, the high proportion of participants with higher income may partly reflect the predominantly Java-based study population rather than a socioeconomic characteristic of contact lens users nationally. With respect to duration of contact lens wear, 42.8% had worn contact lenses for 1–5 years, 27.0% for 6–10 years, 22.4% for 11–20 years, 6.3% for less than 1 year, and 1.4% for more than 20 years (Table 1).

**Table 1.** Characteristics of included participants.

| **Characteristic** | **n** | **%** |
| --- | --- | --- |
| **Age Category** |  |  |
| 18–30 years | 189 | 54.0 |
| 31–40 years | 161 | 46.0 |
| **Gender** |  |  |
| Male | 16 | 4.6 |
| Female | 334 | 95.4 |
| **Education Level** |  |  |
| High school or equivalent | 27 | 7.7 |
| Diploma | 18 | 5.1 |
| Bachelor's degree | 233 | 66.6 |
| Postgraduate degree (Master's/Doctorate) | 72 | 20.6 |
| **Employment Status** |  |  |
| Private-sector employee | 124 | 35.4 |
| Doctor | 67 | 19.1 |
| Student | 46 | 13.1 |
| Housewife | 40 | 11.4 |
| Entrepreneur | 40 | 11.4 |
| Healthcare professional | 13 | 3.7 |
| Teacher | 7 | 2.0 |
| Unemployed | 6 | 1.7 |
| Factory worker/Laborer | 3 | 0.9 |
| Freelance worker | 2 | 0.6 |
| Psychologist | 2 | 0.6 |
| **Marital Status** |  |  |
| Married | 202 | 57.7 |
| Single | 143 | 40.9 |
| Divorced | 5 | 1.4 |
| **Monthly income** |  |  |
| <Rp 3.000.000 | 75 | 21.4 |
| Rp3.000.000-5.000.000 | 48 | 13.7 |
| >Rp5.000.000 | 227 | 64.9 |
| **Region** |  |  |
| Jawa | 269 | 76.9 |
| Sumatra | 49 | 14 |
| Sulawesi | 16 | 4.6 |
| Kalimantan | 10 | 2.9 |
| Papua | 3 | 0.9 |
| Nusa Tenggara and Bali | 2 | 0.6 |
| Maluku | 1 | 0.3 |
| **Duration of contact lens wear** |  |  |
| < 1 year | 22 | 6.3 |
| 1-5 year | 149 | 42.8 |
| 6-10 year | 94 | 27.0 |
| 11-20 year | 78 | 22.4 |
| >20 year | 5 | 1.4 |

### Contact lens wear profile

Soft colored contact lenses were the most frequently used type (60.1%), followed by soft clear contact lenses (30.56%), toric/cylindrical soft lenses (5.05%), lenses of unknown type (2.78%), orthokeratology (Ortho-K) lenses (1.26%), and rigid contact lenses (0.25%). Significant gender differences were observed for soft colored lenses (Fig 1.), which were used by a substantially higher proportion of female (70.7%) than male participants (12.5%; p<0.001), and for soft clear lenses, which were more common among male (81.3%) than female participants (32.3%; p<0.001). No significant gender differences were found for toric, unknown-type, Ortho-K, or rigid lenses. By age group, soft colored lens use was more prevalent among participants aged 18–30 years (75.1%) than those aged 31–40 years (59.6%; p=0.002), whereas soft clear lens use was more common among the 31–40-year group (44.7%) than the 18–30-year group (25.9%; p<0.001). Ortho-K lens use was reported exclusively among participants aged 31–40 years (3.1% vs. 0.0%; p=0.020). No other significant age-related differences were observed (Table 2).

**Figure 1.**
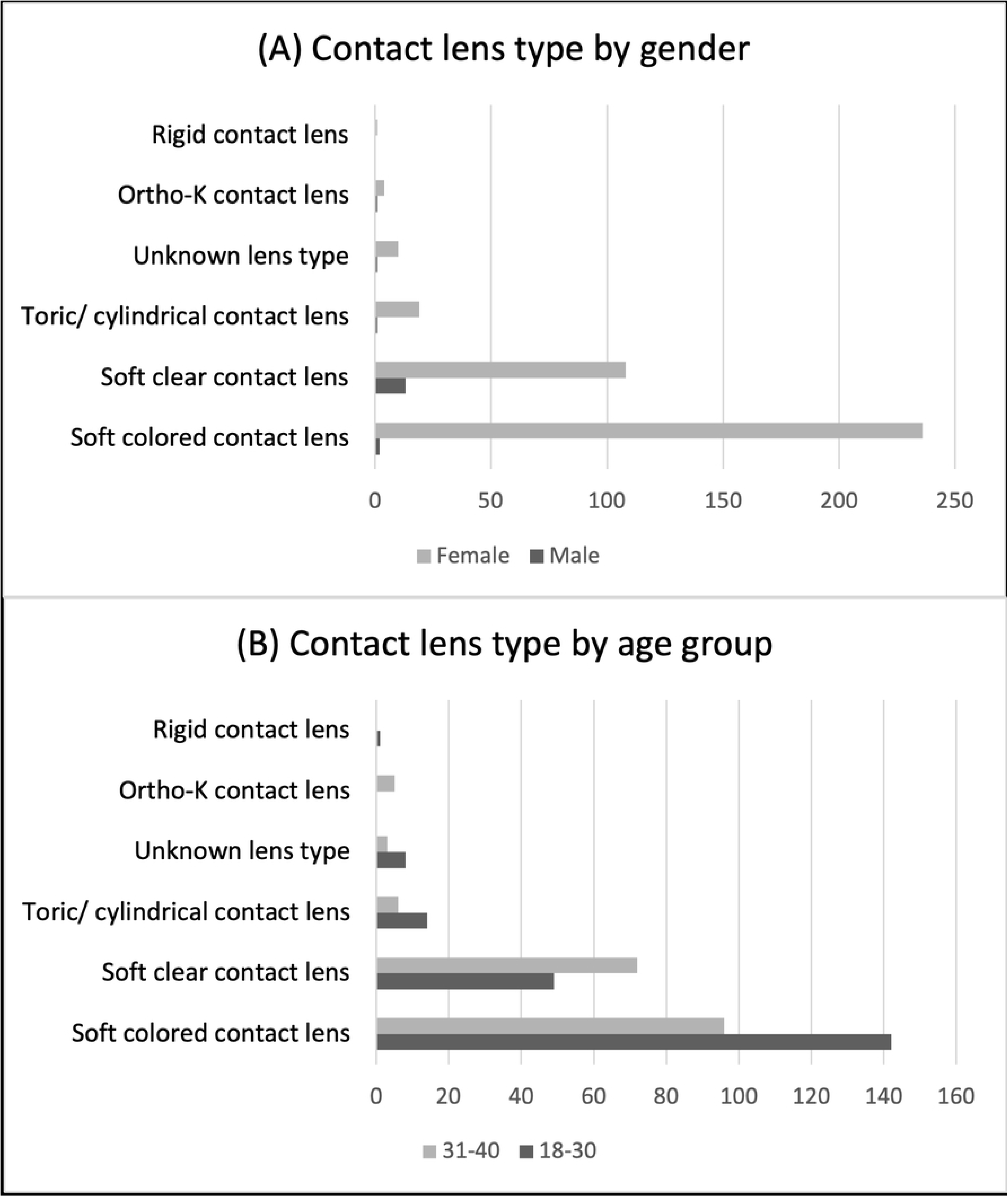
Distribution of contact lens type by (A) age and (B) gender group of wearers.

**Table 2.** Distribution of Contact Lens Types Overall and According to Gender and Age Category.

| Contact Lens Type* | Overall n (%) | Gender<sup>§</sup> |  | p-value† | Age group<sup>§</sup> |  | p-value |
| --- | --- | --- | --- | --- | --- | --- | --- |
|  |  | Male (n=16) n (%) | Female (n=334) n (%) |  | 18–30 years (n=189) n (%) | 31–40 years (n=161) n (%) |  |
| Soft colored contact lens | 238 (60.1) | 2 (12.5) | 236 (70.7) | <0.001 | 142 (75.1) | 96 (59.6) | 0.002 |
| Soft clear contact lens | 121 (30.56) | 13 (81.3) | 108 (32.3) | <0.001 | 49 (25.9) | 72 (44.7) | <0.001 |
| Toric/Cylindrical soft contact lens | 20 (5.05) | 1 (6.3) | 19 (5.7) | 1.000 | 14 (7.4) | 6 (3.7) | 0.169 |
| Unknown lens type | 11 (2.78) | 1 (6.3) | 10 (3.0) | 0.407 | 8 (4.2) | 3 (1.9) | 0.236 |
| Ortho-K contact lens | 5 (1.26) | 1 (6.3) | 4 (1.2) | 0.210 | 0 (0.0) | 5 (3.1) | 0.020 |
| Rigid contact lens | 1 (0.25) | 0 (0.0) | 1 (0.3) | 1.000 | 1 (0.5) | 0 (0.0) | 1.000 |

\*Overall percentages are based on the total number of contact lens type chosen, in which one participant could have more than one type of contact lens (N = 396).
§Gender and age group percentages are based on the number of participants in each group who wore each contact lens type.
**Note.** Data are presented as frequency (n) and percentage (%). Multiple responses were allowed; therefore, percentages do not total 100%. †P-values were obtained using Pearson's chi-square test or Fisher's exact test when expected cell counts were less than five.

Practicality was the most commonly reported reason for contact lens use (38.14%), followed by cosmetic purposes (31.08%) and refractive correction (30.76%). A significant gender difference was found for cosmetic use, which was more frequently reported by female (57.2%) than male participants (18.8%; p=0.003) (Fig 2.). No significant gender differences were observed for refractive correction or practicality, and no significant age-related differences were found for any of the three reasons (Table 3).

**Figure 2.**
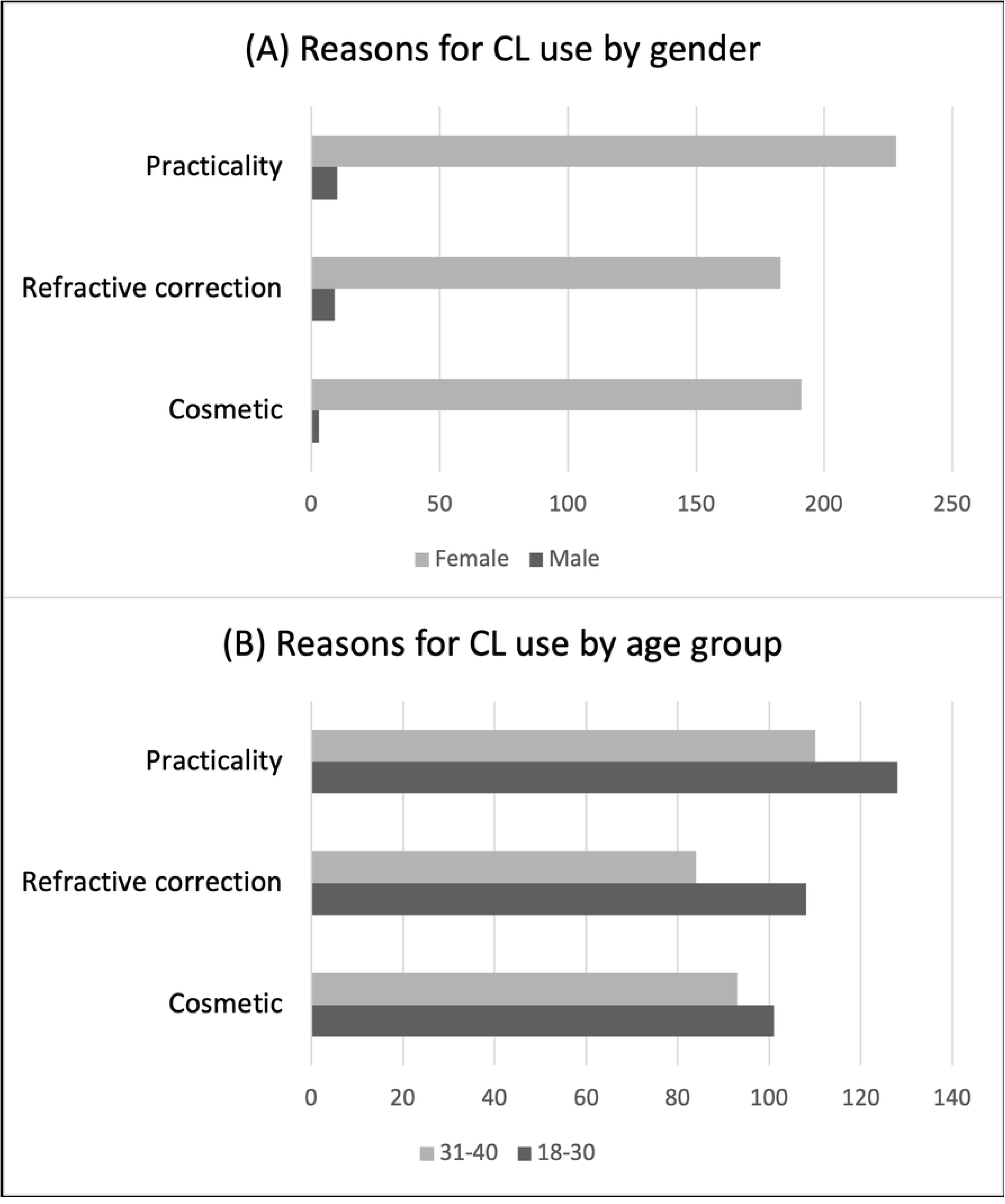
Reasons of contact lens use by (A) age and (B) gender group of wearers.

**Table 3.** Reasons for Contact Lens Use Overall and According to Gender and Age Category.

| Reason for Contact Lens Use | Overall n (%) | Gender<sup>§</sup> |  | p-value† | Age group<sup>§</sup> |  | p-value |
| --- | --- | --- | --- | --- | --- | --- | --- |
|  |  | Male (n=16) n (%) | Female (n=334) n (%) |  | 18–30 years (n=189) n (%) | 31–40 years (n=161) n (%) |  |
| Cosmetic | 194 (31.08) | 3 (18.8) | 191 (57.2) | 0.003 | 101 (53.4) | 93 (57.8) | 0.241 |
| Refractive correction | 192 (30.76) | 9 (56.3) | 183 (54.8) | 0.559 | 108 (57.1) | 84 (52.2) | 0.205 |
| Practicality | 238 (38.14) | 10 (62.5) | 228 (68.3) | 0.406 | 128 (67.7) | 110 (68.3) | 0.499 |

\*Overall percentages are based on the total number of reason for contact lens use, in which one participant could have more than one reason for contact lens use (N = 624).
§Gender and age group percentages are based on the number of participants in each group who answered reasons for contact lens use.
†P-values were obtained using Pearson's chi-square test or Fisher's exact test when expected cell counts were less than five.

### Association between duration of contact lens wear and lens type and reason for use

Among the 348 participants with complete data on lens type and reason for use, no significant association was found between duration of contact lens wear and lens type, with the exception of Ortho-K lens use, which varied significantly across duration categories (p=0.034; Table 4). Similarly, duration of contact lens wear was not significantly associated with cosmetic or refractive correction motives, but was significantly associated with practicality/convenience as a reason for use (p<0.001; Table 5).

**Table 4.** Association between duration of contact lens wear and contact lens type (N = 348)

| Contact lens type | <1 year n (%) | 1–5 years n (%) | 6–10 years n (%) | 11–20 years n (%) | >20 years n (%) | *p*-value† |
| --- | --- | --- | --- | --- | --- | --- |
| Soft colored | 17 (77.3) | 102 (68.5) | 67 (71.3) | 48 (61.5) | 2 (40.0) | 0.328 |
| Soft clear | 4 (18.2) | 47 (31.5) | 32 (34.0) | 36 (46.2) | 2 (40.0) | 0.096 |
| Toric soft | 3 (13.6) | 11 (7.4) | 2 (2.1) | 3 (3.8) | 1 (20.0) | 0.094 |
| Cosmetic lens (unknown type)‡ | 0 (0.0) | 8 (5.4) | 1 (1.1) | 2 (2.6) | 0 (0.0) | 0.320 |
| Ortho-K | 2 (9.1) | 2 (1.3) | 1 (1.1) | 0 (0.0) | 0 (0.0) | **0.034** |
| Rigid contact lens | 0 (0.0) | 1 (0.7) | 0 (0.0) | 0 (0.0) | 0 (0.0) | 0.855 |

† Fisher's exact test.
‡ Replace with the appropriate label if "Unknown lens type" was used as a survey response rather than a true lens category.

**Table 5.**
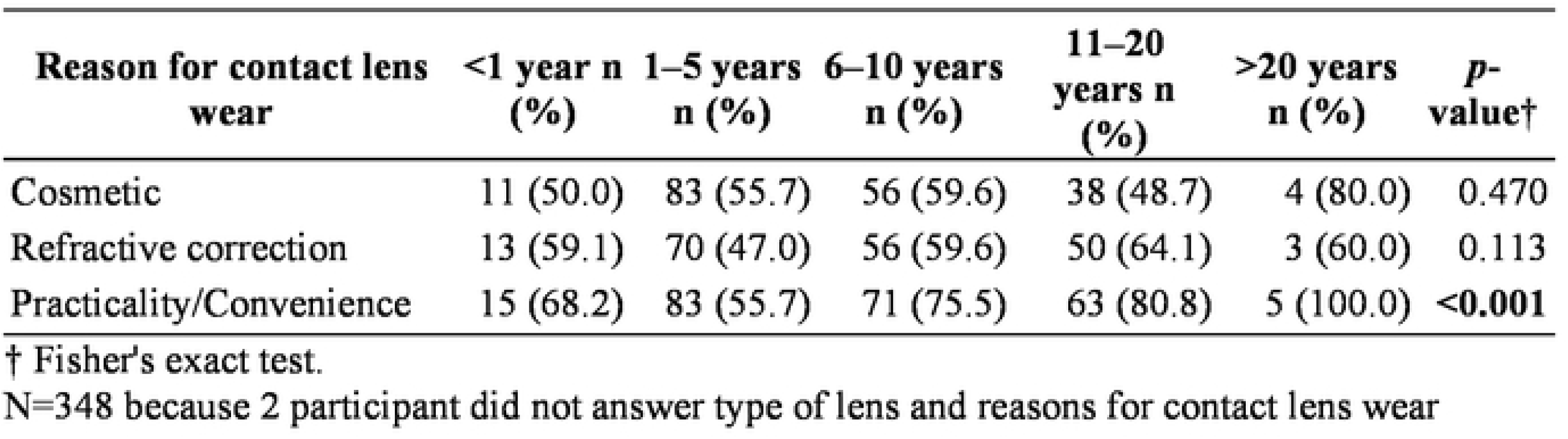
Association between duration of contact lens wear and reason for contact lens wear (N = 348)

### Distribution of refractive errors

Refractive error data were available for 528 eyes (263 right eyes [OD] and 265 left eyes [OS]). Mild myopia was the most common refractive error overall (40.0%), followed by moderate myopia (21.6%). In astigmatic subjects, the majority of contact lens use were by mild astigmatic subjects (14.4%). The distribution was similar between the right and left eyes (Table 6.).

**Table 6.** Distribution of Refractive Errors by Eye and Severity (N = 528 Eyes)

| **Refractive Error** | **OD (n = 263)** | **OS (n = 265)** | **Total Eyes (n = 528)** |
| --- | --- | --- | --- |
| Emmetropia | 22 (8.4%) | 21 (7.9%) | 43 (8.1%) |
| Mild astigmatism | 34 (12.9%) | 42 (15.8%) | 76 (14.4%) |
| Moderate astigmatism | 14 (5.3%) | 10 (3.8%) | 24 (4.5%) |
| High astigmatism | 1 (0.4%) | 1 (0.4%) | 2 (0.4%) |
| Mild myopia | 106 (40.3%) | 105 (39.6%) | 211 (40.0%) |
| Moderate myopia | 56 (21.3%) | 58 (21.9%) | 114 (21.6%) |
| High myopia | 26 (9.9%) | 24 (9.1%) | 50 (9.5%) |
| Hyperopia | 4 (1.5%) | 4 (1.5%) | 8 (1.5%) |
| **Total** | **263 (100.0%)** | **265 (100.0%)** | **528 (100.0%)** |

Percentages were calculated using the number of eyes with complete refractive data (OD = 263, OS = 265, total = 528). Eyes with missing or incomplete refractive error data (e.g., missing positive [+] or negative [-] signs, or non-uniform notation between eyes), or if no refractive error measurement was reported (n = 172) were excluded from this analysis.

### Contact lens correction status and agreement between recommended and actual spherical power

Among 254 eyes, undercorrection was the most frequent contact lens (CL) correction status in both eyes, occurring in 113 eyes (44.4%) in the right eye (OD) and 114 eyes (44.8%) in the left eye (OS). Appropriate correction was observed in 97 eyes (38.2%) for OD and 100 eyes (39.3%) for OS, whereas overcorrection occurred in 44 eyes (17.3%) and 40 eyes (15.7%), respectively (Table 7.).

**Table 7.** Distribution of Contact Lens Correction Status (N = 254)

| **Correction status** | **OD, n (%)** | **OS, n (%)** |
| --- | --- | --- |
| Appropriate | 97 (38.2) | 100 (39.3) |
| Overcorrection | 44 (17.3) | 40 (15.7) |
| Undercorrection | 113 (44.4) | 114 (44.8) |
| **Total** | **254 (100.0)** | **254 (100.0)** |

The recommended CL spherical power was more negative than the actual CL power in both eyes. For OD, the mean recommended power was −3.17 ± 2.05 D compared with an actual power of −2.99 ± 2.20 D, yielding a mean difference of −0.18 ± 1.19 D (95% CI, −0.33 to −0.04; *p* = 0.015). Similarly, for OS, the mean recommended power was −3.04 ± 2.05 D compared with an actual power of −2.80 ± 2.06 D, with a mean difference of −0.24 ± 1.16 D (95% CI, −0.39 to −0.10; *p* = 0.001). Thus, the actual CL power was significantly less negative than the recommended power in both eyes, indicating a systematic tendency toward undercorrection (Table 8.).

**Table 8.** Difference Between Recommended and Actual Contact Lens Spherical Power (N = 254)

| **Eye** | **Recommended CL power, Mean ± SD (D)** | **Actual CL power, Mean ± SD (D)** | **Mean difference ± SD (D)** | **95% CI (D)** | **p value** |
| --- | --- | --- | --- | --- | --- |
| **OD** | -3.17 ± 2.05 | -2.99 ± 2.20 | -0.18 ± 1.19 | -0.33 to -0.04 | **0.015** |
| **OS** | -3.04 ± 2.05 | -2.80 ± 2.06 | -0.24 ± 1.16 | -0.39 to -0.10 | **0.001** |

### Factors associated with contact lens undercorrection

Multinomial logistic regression showed that myopia severity was the only factor significantly associated with CL correction status in both eyes. Compared with appropriate correction, greater myopia severity was strongly associated with undercorrection in both OD (OR 5.01, 95% CI 2.88– 8.74; *p* < 0.001) and OS (OR 3.63, 95% CI 2.20–5.99; *p* < 0.001). Myopia severity was also significantly associated with overcorrection in OD (OR 5.83, 95% CI 3.06–11.10; *p* < 0.001) and OS (OR 4.22, 95% CI 2.30–7.73; *p* < 0.001).

In contrast, age, duration of CL wear, and sex were not significantly associated with either undercorrection or overcorrection in either eye (Table 9.). For undercorrection, age showed ORs of 0.94 (*p* = 0.834) for OD and 1.28 (*p* = 0.418) for OS, while duration of CL wear showed ORs of 0.95 (*p* = 0.795) and 0.91 (*p* = 0.584), respectively. Male sex was not significantly associated with undercorrection in either eye (OD: OR 0.16, 95% CI 0.03–1.01, *p* = 0.051; OS: OR 0.71, 95% CI 0.18–2.77, *p* = 0.616).

**Table 9.** Multinomial Logistic Regression Analysis of Factors Associated with Contact Lens Correction Status.

| Variable | OD |  | OD |  | OS |  | OS |  |
| --- | --- | --- | --- | --- | --- | --- | --- | --- |
|  | Overcorrection<br>OR (95% CI) | *p* | Undercorrection<br>OR (95% CI) | *p* | Overcorrection<br>OR (95% CI) | *p* | Undercorrection<br>OR (95% CI) | *p* |
| Age | 0.60 (0.26–1.38) | 0.231 | 0.94 (0.50–1.75) | 0.834 | 0.96 (0.42–2.17) | 0.919 | 1.28 (0.70–2.33) | 0.418 |
| Duration<br>of contact<br>lens wear | 1.48 (0.93–2.35) | 0.100 | 0.95 (0.67–1.36) | 0.795 | 1.21 (0.76–1.91) | 0.424 | 0.91 (0.65–1.28) | 0.584 |
| Myopia<br>severity | **5.83 (3.06–**<br>**11.10)** | **<0.001** | **5.01 (2.88–8.74)** | **<0.001** | **4.22 (2.30–7.73)** | **<0.001** | **3.63 (2.20–5.99)** | **<0.001** |
| Male sex | 1.02 (0.19–5.47) | 0.977 | 0.16 (0.03–1.01) | 0.051 | 0.35 (0.04–3.56) | 0.375 | 0.71 (0.18–2.77) | 0.616 |

*Reference category for the dependent variable: appropriate. Female sex was the reference category for sex. Age 18–30 as the reference. Duration of CL wear < 1 year as the reference.*

## Discussion

This cross-sectional study evaluated the characteristics of contact lens wearer in Indonesia, their correction status, and factors associated with its correction status. The majority of participants were employed, married, female between 18-30 years of age with a bachelor degree and monthly income of more than five million rupiah.

Data on age of contact lens prescription from 20 countries reported similar average age of contact lens prescription from this study (29.52 ± 4.31), which were 30.8 ± 13.9 years and 32.5 ± 14.3 years for males and females, respectively.^[22]^ Moreover, the average age were also similar with the mean age from 100 countries reported in 2025, being 34.7 ± 15.5.^[10]^ The findings that most CL wearers were females aligned with results from previous demographic studies.^[7,9,23–25]^ Most previous studies reported that fashion or cosmetic were the most common reasons for CL use amongst females ^[9,23]^, in the contrary, this study reported that practicality as the most common reason. This may be supported by studies reporting that contact lens wear has favorable findings regarding visual related quality of life compared to spectacle wear.^[26]^ However, it must also be acknowledged that the majority of participants wore soft-colored contact lens (60.1%), therefore the assumption that cosmetic or fashion reasons were one of the deciding factors of contact lens wear should still be highlighted. Aligned with the results of this study, the 2025 report on contact lens prescribing reported that soft-contact lens comprises the majority (88%) of CL prescribed in 100 countries.^[10]^

The majority of participants in this study have worn CL for 1-5 years (42.8%), and the duration of wear did not differ significantly between different contact lens type except for orthokeratology, as orthokeratology had only been prescribed in Indonesia for the last 10-20 years.^[24]^ Moreover, the low percentage of ortho-K use also aligned with the 2025 international prescription report (2%).^[14]^

Mild myopia comprises as the most prevalent (40%) refractive error in our study. Undercorrection was the most common contact lens (CL) correction status in both eyes, accounting for 44.4% of OD and 44.8% of OS, whereas appropriate correction was observed in 38.2% and 39.3%, respectively. There has been no previous study reporting reasons for over or undercorrection of contact lens prescription. However, inappropriate contact lens use may be contributed by the accessibility of contact lens purchase in beauty salons and pharmacies without prescription by an ophthalmologist or optometrist.^[9,25,27]^

The actual CL spherical power differed significantly from the recommended power in both OD (mean difference −0.18 D; p=0.015) and OS (−0.24 D; p=0.001). Although these differences reached statistical significance, their clinical significance may be more limited. The average discrepancy was 0.18 D in OD and 0.24 D in OS, which is relatively small compared with the overall variability in prescription differences. Importantly, the mean difference does not indicate that every patient experienced a clinically meaningful discrepancy; rather, it reflects the average difference across the population.

Greater myopia severity was significantly associated with both overcorrection and undercorrection in both eyes, with ORs ranging from 3.63 to 5.83 (all p<0.001). Age, duration of CL wear, and sex were not significantly associated with correction status. These findings suggest that increasing myopia severity is an important factor associated with CL prescription mismatch, although it does not predict the direction of the mismatch. Inappropriate myopia correction has been reported to enhance myopia progression in children^[28–31]^ and adults.^[32]^ However, it remains unclear whether the undercorrection observed in this study reflects an intentional choice by the wearer or an unintended outcome of the prescribing process. One plausible explanation is that both overcorrection and undercorrection stem from gaps in prescription-related knowledge that are not necessarily attributable to the eye care practitioner (ECP); many contact lens users may be unaware that vertex distance needs to be accounted for when converting a spectacle prescription to a contact lens prescription. Therefore, the findings in our study which reported greater myopia severity as factors associated with inappropriate correction presents as a clinically relevant problem in contact lens prescribing and purchase in Indonesia.

### Limitations and strengths

This study has several limitations. First, its cross-sectional design and convenience sampling of volunteer participants may limit the generalizability of the findings. In addition, refractive error and contact lens prescription data were self-reported rather than obtained from clinical records or refractive assessment, which may introduce recall and reporting bias. Although responses with uninterpretable refractive values were excluded, the distribution and severity of refractive errors should therefore be interpreted with caution. Second, the sample was predominantly composed of participants residing in Java (76.9%), with substantially smaller representation from other regions of Indonesia. This may limit the representativeness of the findings for the Indonesian population as a whole, particularly because access to eye-care services, contact lens availability, socioeconomic conditions, and patterns of technology adoption may differ across regions. Third, the study did not evaluate habitual CL replacement schedule (daily, biweekly, monthly, or yearly disposable), duration of CL wear, as well as contact lens care practices, including cleaning, disinfection, storage, replacement, and hygiene behaviors. Consequently, the study provides limited information regarding the relationship between demographic or prescription characteristics and the safety of contact lens wear. Despite these limitations, to our knowledge, this study represents the first nationwide demographic study specifically describing contact lens wearers in Indonesia. The study provides a broad overview of contact lens wearer characteristics, lens types, reasons for use, refractive error distribution, and correction status, while also examining factors associated with prescription mismatch. These findings provide an important baseline for understanding contact lens use in Indonesia and may help inform future population-based studies and strategies to improve contact lens prescribing and safe use.

## Conclusion

This study provides a demographic profile of contact lens users and identifies patterns relevant to public education on appropriate contact lens use. Colored soft contact lenses were the most commonly purchased type. Although adverse events were not assessed in this study, the high use of colored lenses highlights an important target for education, particularly regarding appropriate prescription, fitting, purchasing, lens care, and potential complication.

Public education should emphasize that contact lenses, including colored lenses, should be selected and fitted by an eye-care professional rather than purchased solely based on cosmetic preference or availability. Users should also be educated regarding proper lens hygiene, replacement schedules, and warning symptoms requiring prompt eye examination. Further studies should evaluate contact lens purchasing practices and associated ocular complications in Indonesian users.

## DECLARATIONS

If any of these sections are not relevant to your manuscript, please include the heading and write ‘Not applicable’ for that section.

Funding: None

Conflicts of interest/Competing interests: None Availability of data and material: Available

Ethics approval (Title of Ethics Committee, Country, and approval code): Ethics Committee of the Faculty of Medicine, the University of Indonesia, Indonesia (protocol number: 1196/UN2.F1/ETIK/PPM.00.01/2024).

## Data Availability

The dataset will be available upon publication through supplementary materials.

## Reference

1. Gammoh Y, Abdu M. Contact lens procurement and usage habits among adults in Sudan. PLOS ONE. 2021;16(5):e0251987. doi:10.1371/journal.pone.0251987

2. Stapleton F, Tan J. Impact of Contact Lens Material, Design, and Fitting on Discomfort. Eye Contact Lens. 2017;43(1):32–9. doi:10.1097/ICL.0000000000000318 PubMed PMID: 28002225.

3. Gurnani B, Kaur K. Contact Lenses. In: StatPearls [Internet]. Treasure Island (FL): StatPearls Publishing; 2026 [cited 2026 Jun 26]. Available from: http://www.ncbi.nlm.nih.gov/books/NBK580554/ PubMed PMID: 35593861.

4. Dumbleton K, Caffery B, Dogru M, Hickson-Curran S, Kern J, Kojima T, et al. The TFOS International Workshop on Contact Lens Discomfort: report of the subcommittee on epidemiology. Invest Ophthalmol Vis Sci. 2013;54(11):TFOS20-36. doi:10.1167/iovs.13-13125 PubMed PMID: 24058130.

5. Morgan PB, Efron N, Woods CA, Santodomingo-Rubido J, International Contact Lens Prescribing Survey Consortium. International survey of orthokeratology contact lens fitting. Contact Lens Anterior Eye J Br Contact Lens Assoc. 2019;42(4):450–4. doi:10.1016/j.clae.2018.11.005 PubMed PMID: 30448008.

6. Li W, Sun X, Wang Z, Zhang Y. A survey of contact lens-related complications in a tertiary hospital in China. Contact Lens Anterior Eye J Br Contact Lens Assoc. 2018;41(2):201–4. doi:10.1016/j.clae.2017.10.007 PubMed PMID: 29033270.

7. Ocansey S, Ovenseri Ogbomo G, Abu EK, Morny EKA, Adjei-Boye O. Profile, knowledge, and attitude of contact lens users regarding contact lens wear in Ghana. Contact Lens Anterior Eye J Br Contact Lens Assoc. 2019;42(2):170–7. doi:10.1016/j.clae.2018.10.012 PubMed PMID: 30415960.

8. Lee YC, Lim CW, Saw SM, Koh D. The prevalence and pattern of contact lens use in a Singapore community. CLAO J Off Publ Contact Lens Assoc Ophthalmol Inc. 2000;26(1):21–5. PubMed PMID: 10656305.

9. Abahussin M, AlAnazi M, Ogbuehi KC, Osuagwu UL. Prevalence, use and sale of contact lenses in Saudi Arabia: survey on university women and non-ophthalmic stores. Contact Lens Anterior Eye J Br Contact Lens Assoc. 2014;37(3):185–90. doi:10.1016/j.clae.2013.10.001 PubMed PMID: 24211011.

10. PentaVision [Internet]. [cited 2026 Aug 24]. International Contact Lens Prescribing in 2025. Available from: https://clspectrum.com/issues/2026/january-february/international-contact-lens-prescribing-in-2025

11. Morgan PB, Efron N, Helland M, Itoi M, Jones D, Nichols JJ, et al. Global trends in prescribing contact lenses for extended wear. Contact Lens Anterior Eye J Br Contact Lens Assoc. 2011;34(1):32–5. doi:10.1016/j.clae.2010.06.007 PubMed PMID: 20630794.

12. Morgan PB, Efron N, Woods CA, International Contact Lens Prescribing Survey Consortium. Determinants of the frequency of contact lens wear. Eye Contact Lens. 2013;39(3):200–4. doi:10.1097/ICL.0b013e31827a7ad3 PubMed PMID: 23629005.

13. Chalmers RL, Hunt C, Hickson-Curran S, Young G. Struggle with hydrogel CL wear increases with age in young adults. Contact Lens Anterior Eye J Br Contact Lens Assoc. 2009;32(3):113–9. doi:10.1016/j.clae.2008.12.001 PubMed PMID: 19201645.

14. Efron N, Morgan PB, Woods CA, Jones D, Jones L, Santodomingo-Rubido J, et al. International trends in prescribing contact lenses for myopia control (2011–2024): An update. Contact Lens Anterior Eye. 2025;48(5):102451. doi:10.1016/j.clae.2025.102451

15. Lee JRJ, Yee TH, Levitz D, Lim BXH, Mehta JS, Stapleton F, et al. A Review of Contact Lens Regulations in the Asia Pacific Region. Eye Contact Lens. 2025;51(3):e149–56. doi:10.1097/ICL.0000000000001150 PubMed PMID: 39642261.

16. Liang J, Pu Y, Chen J, Liu M, Ouyang B, Jin Z, et al. Global prevalence, trend and projection of myopia in children and adolescents from 1990 to 2050: a comprehensive systematic review and meta-analysis. Br J Ophthalmol. 2025;109(3):362–71. doi:10.1136/bjo-2024-325427 PubMed PMID: 39317432.

17. Mahayana IT, Indrawati SG, Pawiroranu S. The prevalence of uncorrected refractive error in urban, suburban, exurban and rural primary school children in Indonesian population. Int J Ophthalmol. 2017;10(11):1771–6. doi:10.18240/ijo.2017.11.21 PubMed PMID: 29181324; PubMed Central PMCID: PMC5686379.

18. Flitcroft DI, He M, Jonas JB, Jong M, Naidoo K, Ohno-Matsui K, et al. IMI – Defining and Classifying Myopia: A Proposed Set of Standards for Clinical and Epidemiologic Studies. Invest Ophthalmol Vis Sci. 2019;60(3):M20–30. doi:10.1167/iovs.18-25957 PubMed PMID: 30817826; PubMed Central PMCID: PMC6735818.

19. Zhang J, Wu Y, Sharma B, Gupta R, Jawla S, Bullimore MA. Epidemiology and Burden of Astigmatism: A Systematic Literature Review. Optom Vis Sci. 2023;100(3):218–31. doi:10.1097/OPX.0000000000001998 PubMed PMID: 36749017; PubMed Central PMCID: PMC10045990.

20. Vertex distance - American Academy of Ophthalmology [Internet]. [cited 2026 Aug 21]. Available from: https://www.aao.org/education/image/vertex-distance-in-correction-of-high-refractive-e

21. Inilah Daftar UMR 2026 di Indonesia & Rata-rata Kenaikannya [Internet]. [cited 2026 Aug 26]. Available from: https://pegadaian.co.id/artikel/keuangan/daftar-umr-2026-resmi

22. Morgan PB, Efron N. Global contact lens prescribing 2000-2020. Clin Exp Optom. 2022;105(3):298–312. doi:10.1080/08164622.2022.2033604 PubMed PMID: 35184672.

23. Ezinne NE, Bhattarai D, Ekemiri KK, Harbajan GN, Crooks AC, Mashige KP, et al. Demographic profiles of contact lens wearers and their association with lens wear characteristics in Trinidad and Tobago: A retrospective study. PloS One. 2022;17(7):e0264659. doi:10.1371/journal.pone.0264659 PubMed PMID: 35867670; PubMed Central PMCID: PMC9307171.

24. Hwang SD, Jung WY, Lee S, Kim SR, Park M. Sociodemographic and refractive factors associated with contact lens use: Insights from a nationwide Korean survey. Contact Lens Anterior Eye J Br Contact Lens Assoc. 2026;49(3):102634. doi:10.1016/j.clae.2026.102634 PubMed PMID: 41844082.

25. Kim M, Paik JS, Kim D, Hwang HS, Han K, Na KS. Current status of contact lenses usage in Korea: A population-based cohort study 2021. PLOS ONE. 2024;19(3):e0296279. doi:10.1371/journal.pone.0296279 PubMed PMID: 38507419; PubMed Central PMCID: PMC10954094.

26. Osae EA, Osei KA, Edwards NC, Wygonik E, Blackie CA. Contact Lenses and Vision-Related Quality of Life: Evidence and Clinical Insights. Clin Optom. 2026;18:598390. doi:10.2147/OPTO.S598390 PubMed PMID: 42465553; PubMed Central PMCID: PMC13374603.

27. Fogel J, Zidile C. Contact lenses purchased over the internet place individuals potentially at risk for harmful eye care practices. Optometry. 2008;79(1):23–35. doi:10.1016/j.optm.2007.07.013 PubMed PMID: 18156093.

28. Yazdani N, Sadeghi R, Ehsaei A, Taghipour A, Hasanzadeh S, Zarifmahmoudi L, et al. Under-correction or full correction of myopia? A meta-analysis. J Optom. 2021;14(1):11–9. doi:10.1016/j.optom.2020.04.003 PubMed PMID: 32507615; PubMed Central PMCID: PMC7752985.

29. Logan NS, Wolffsohn JS. Role of un-correction, under-correction and over-correction of myopia as a strategy for slowing myopic progression. Clin Exp Optom. 2020;103(2):133–7. doi:10.1111/cxo.12978 PubMed PMID: 31854025.

30. Atchison DA, Schmid KL, Edwards KP, Muller SM, Robotham J. The effect of under and over refractive correction on visual performance and spectacle lens acceptance. Ophthalmic Physiol Opt J Br Coll Ophthalmic Opt. 2001;21(4):255–61. doi:10.1046/j.1475-1313.2001.00588.x PubMed PMID: 11430618.

31. Chung K, Mohidin N, O’Leary DJ. Undercorrection of myopia enhances rather than inhibits myopia progression. Vision Res. 2002;42(22):2555–9. doi:10.1016/S0042-6989(02)00258-4

32. Vasudevan B, Esposito C, Peterson C, Coronado C, Ciuffreda KJ. Under-correction of human myopia--is it myopigenic?: a retrospective analysis of clinical refraction data. J Optom. 2014;7(3):147–52. doi:10.1016/j.optom.2013.12.007 PubMed PMID: 25000870; PubMed Central PMCID: PMC4087177.

